# Epidemics, Air Travel, and Elimination in a Globalized World: The Case of Measles

**DOI:** 10.1101/2020.05.08.20095414

**Authors:** Shaun Truelove, Luis Mier-y-Teran-Romero, Paul Gastanaduy, Allison Taylor Walker, Andre Berro, Justin Lessler, Michael A. Johansson

## Abstract

Although the United States (U.S.) declared measles eliminated in 2000, so long as measles circulates globally, continued importations will trigger sporadic U.S. outbreaks. The United States is highly connected to the rest of the world, and importations occur largely as a result of air travel by U.S. residents and international visitors. We developed a model to assess the risk of measles virus importation from Europe, Asia, and Africa through air travel. We projected 308 (95% prediction interval, 151-518) total measles cases imported from 66 countries during 2006-2015; 290 importations were reported. The model projected a spike in importation risk from Israel during October 2018, coincident with large, importation-triggered outbreaks in New York and New Jersey. Our model shows U.S. importation risk primarily originates from European and Asian countries. Models such as this inform preemptive actions to mitigate infectious disease threats, but only if we prioritize global surveillance and data sharing.

## INTRODUCTION

Approximately 78 million tourists^1^ and 44 million residents^2^ enter the United States (U.S.) from abroad via commercial flights every year – equivalent to over 300,000 travelers daily. With each traveler comes the potential for an infectious pathogen to be brought into the United States. The rapid, long-distance transmission potential posed by global air travel has become more evident during the last two decades, particularly in the context of emerging pathogens. In 2002, SARS coronavirus emerged in China. It spread rapidly to 36 countries, transmitted largely by air travelers, causing 8,098 cases and 774 deaths worldwide^3^. Six years later, the 2009 H1N1 influenza pandemic spread quickly across the world from Mexico; it caused 284,000 deaths worldwide and stoked widespread fear of a repeat 1918 influenza pandemic^4^. Recent deadly outbreaks of Middle East respiratory syndrome (MERS) and Ebola have prompted travel advisories, airport screening, and travel restrictions for exposed individuals^5–10^.

While these outbreaks highlight concerns about the spread of emerging pathogens, global travel is similarly critical to the spread and reintroduction of infectious diseases that countries have struggled to eliminate, particularly measles. In 2000, the United States declared measles elimination^11^. However, since 2000, the United States has reported more than 100 measles outbreaks, over 2,000 reported cases, and 3 measles-related deaths, all arising from infections in travelers^12–14^. With increasing globalization, the threat posed by geographically distant outbreaks will continue to grow. Achieving elimination of a highly contagious disease like measles requires immense efforts and resources, and because of the constant threat of reintroduction, maintaining elimination can be challenging and expensive.

Having a model to help predict when and where measles importation may occur, therefore, can assist in planning for resource allocation. Relying on global measles incidence data and 2006-2015 airline itinerary information for international arrivals into the United States from other countries, we quantified the potential for measles importation using a framework previously developed to model arbovirus spread^15,16^. For simplicity, we broadly define this potential as the projected “risk of importation” and quantify it with crude estimates of the absolute number of importations per time period and the ratio of these importations between locations and time periods. Because a multitude of factors can modify how this potential for importation translates into actual imported cases, we focus on this projected potential and not the absolute estimates. We validated our approach with data on reported measles importations. This model provides the risk of measles importation, both where importations are most likely to occur (i.e., into which U.S. cities) and where they are most likely to originate (i.e., from which countries). This work provides a platform that public health officials can use for real-time risk estimates and demonstrates the value of global disease surveillance efforts.

## RESULTS

We estimated the monthly risk of measles importation into the United States from 2006 through 2015 from 66 countries in Europe, Africa, and Asia for which data were available on measles incidence and air travel to the United States. For some countries measles surveillance data were not available for all periods. Hence, we restricted our analysis for each country to months when data were available. The final data set comprised 7,260 country-months (92% of those possible over the study period). During this time, the U.S. Centers for Disease Control and Prevention (CDC) reported 412 confirmed imported measles cases. Of these, 376 (91%) originated in one of the 66 countries included in our analysis, and 290 (70% of all importations) were reported during months when measles surveillance data were available from origin countries. We estimated importation risk for all 7,260 country-months with surveillance data and compared model estimates to the 290 imported cases reported during those months.

Between 2006-2015, the 66 included countries reported 1,320,171 confirmed cases of measles, and travelers made an estimated 340 million trips to the United States from these countries. Global incidence of measles was characterized by annual periodicity, with several distinct outbreaks in Europe, Asia, and Africa (Figure 1). In Africa, large outbreaks occurred in Malawi (134,000 cases; 2010), Nigeria (58,000 cases; 2013), Burkina Faso (54,000 cases; 2009), and Zambia (38,000 cases; 2010-11)^17–20^. In Asia, a large measles outbreak occurred in the Philippines (59,000 cases; 2014), and both India and China reported substantial average annual case counts (34,000 and 47,000, respectively)^21^. In Europe, notable outbreaks occurred in Bulgaria (23,500 cases; 2009-10), France (16,000 cases; 2010-11), and Ukraine (2005-06: 42,000 cases; 2011-12: 14,000 cases)^21–23^. Overall, most travel to the United States originated from Europe (68%) and Asia (30%), with only 2% from Africa. In the most recent years, 2010-2015, travel volume increased consistently, by 4%, 6%, and 10% per year from Europe, Asia, and Africa, respectively (Figure 2).

**Figure 1.**
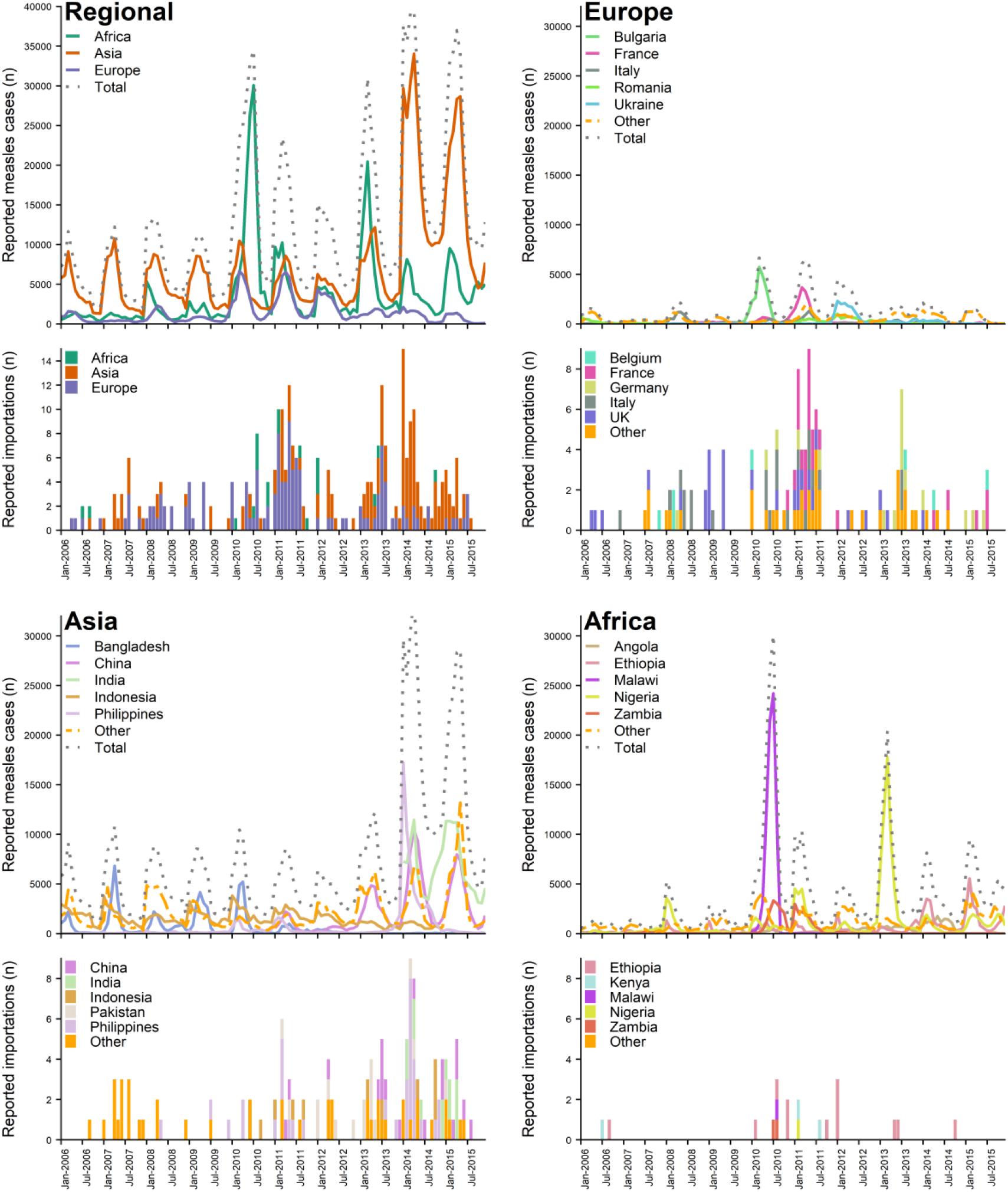
Number of reported measles cases in 66 countries in Europe, Asia, and Africa and importation of cases from these countries into the United States, during January 2006 – December 2015. Reported cases and importations included reflect months when data were complete for the country. For Asia, measles case data were not available for India during 2006-2013 and China 2006-2010, thus reported importations for those countries are not represented in this figure; the jump in reported measles cases in 2014-2015 can be explained by inclusion of India.

**Figure 2.**
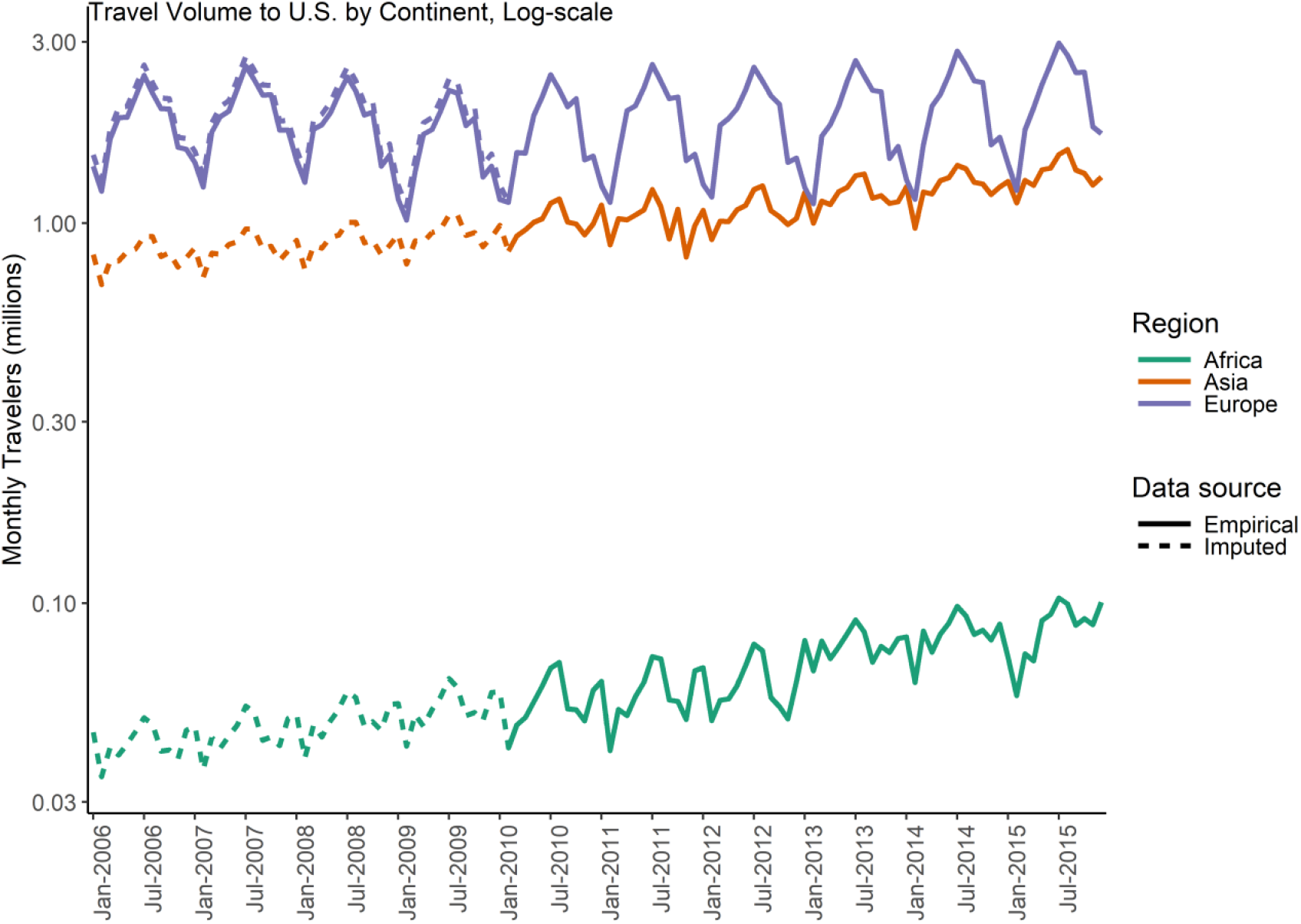
Monthly international air travel volume into the United States, by region, from sixty-six countries during January 2006 – December 2015. Data for Asia, Africa, and six European countries (Belarus, Bosnia & Herzegovina, Croatia, Russia, Switzerland, and Ukraine) were only available for January 2010 – December 2015 (solid lines). Country-specific travel volume data that were unavailable were estimated using available data for 2010-2015 and regional data for the full time period (dotted lines, see Methods).

### Reported measles cases and importations

Reported monthly measles case counts in source countries correlated moderately with reported monthly measles importations into the United States (*r*=0.52; Figure 1). Correlation was highest for Asia (*r*=0.67), where 802,535 reported measles cases resulted in 135 reported importations (Figure 1B, Table1). The number of importations from Europe was similar (n=136), though reported cases (162,779) was lower, as was correlation (*r*=0.39). Despite 362,182 total cases reported from twelve African countries, only 19 importations into the United States were reported, and correlation between reported cases and importations from Africa was low (*r*=0.23).

**Table 1.**
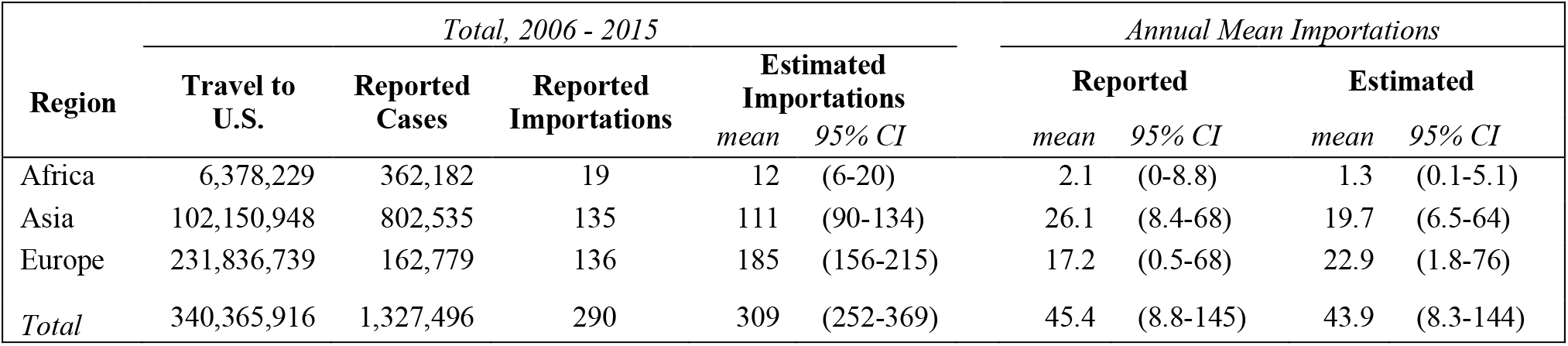
Reported and model estimated total and mean annual number of measles importations into the United States from Europe, Asia, and Africa, during 2006-2015 from 66 countries. Estimated importations were produced by a model that used a combination of reported measles incidence in source countries, source country population, and air travel volume from source countries to the United Stated. Reported data and estimated importations include the 7,200 country-months for which complete measles case and travel data were available. Both reported and estimated annual importations were adjusted for missing data, producing the discrepancies between total and annual mean importations. This was particularly important due to missing case data for China for 2006-2010 and for India for 2006-2013, during which time importations could not be estimated, but 10 and 50 direct importations were reported, respectively.

### Estimated risk of importation into the United States, by source region

To examine contributions by region and country, we compared mean annual estimates of importation and relative risk. Mean annual estimates were used rather than cumulative estimates as data were not available during part of the study period for several high-risk countries (e.g., data from China were not available for 2006-2010, nor were data available from India for 2006-2013, during which 10 and 50 importations occurred respectively). On average, estimated measles importations predominantly originated from Asia and Europe (60% and 35%, respectively), with only 5% reported from Africa annually. Taken alone, neither case nor travel data fully captures of the risk of measles importation: among the three regions, Asia and Africa reported the majority of measles cases (61% and 27%), with only 12% occurring in Europe. In contrast, 68% of travel volume to the United States originated from Europe, compared to 30% from Asia and only 2% from Africa. Using this model and accounts for reported cases, travel volume, and source country population, we estimated an average of 22.9 (95% CI, 1.8-76) measles cases imported from Europe annually and 19.7 (95% CI, 6.5-64) from Asia (Table 1), with Africa contributing just over one measles importation each year (mean=1.3; 95% CI, 0.1-5.1). These are comparable to average annual reported importations of 17.2 (95% CI, 0.5-68) for Europe, 26.1 (95% CI, 8.4-68) for Asia, and 2.1 (95% CI, 0-8.8) for Africa.

### Estimated risk of importation into the United States, by country

The five countries with the highest average annual estimated risk of measles case exportation to the United States were India, the United Kingdom (U.K.), France, the Philippines, and China (Table 2, Figure 3). Risk was greatest for countries with high travel volume to the United States and either persistent measles cases (e.g., the U.K., India, China) or large outbreaks (e.g., France, the Philippines). Yearly country-specific mean importation risk correlated moderately with mean travel volume (*r*=0.42) and reported cases (*r*=0.43), and poorly with population size (*r*=0.20).

**Table 2.**
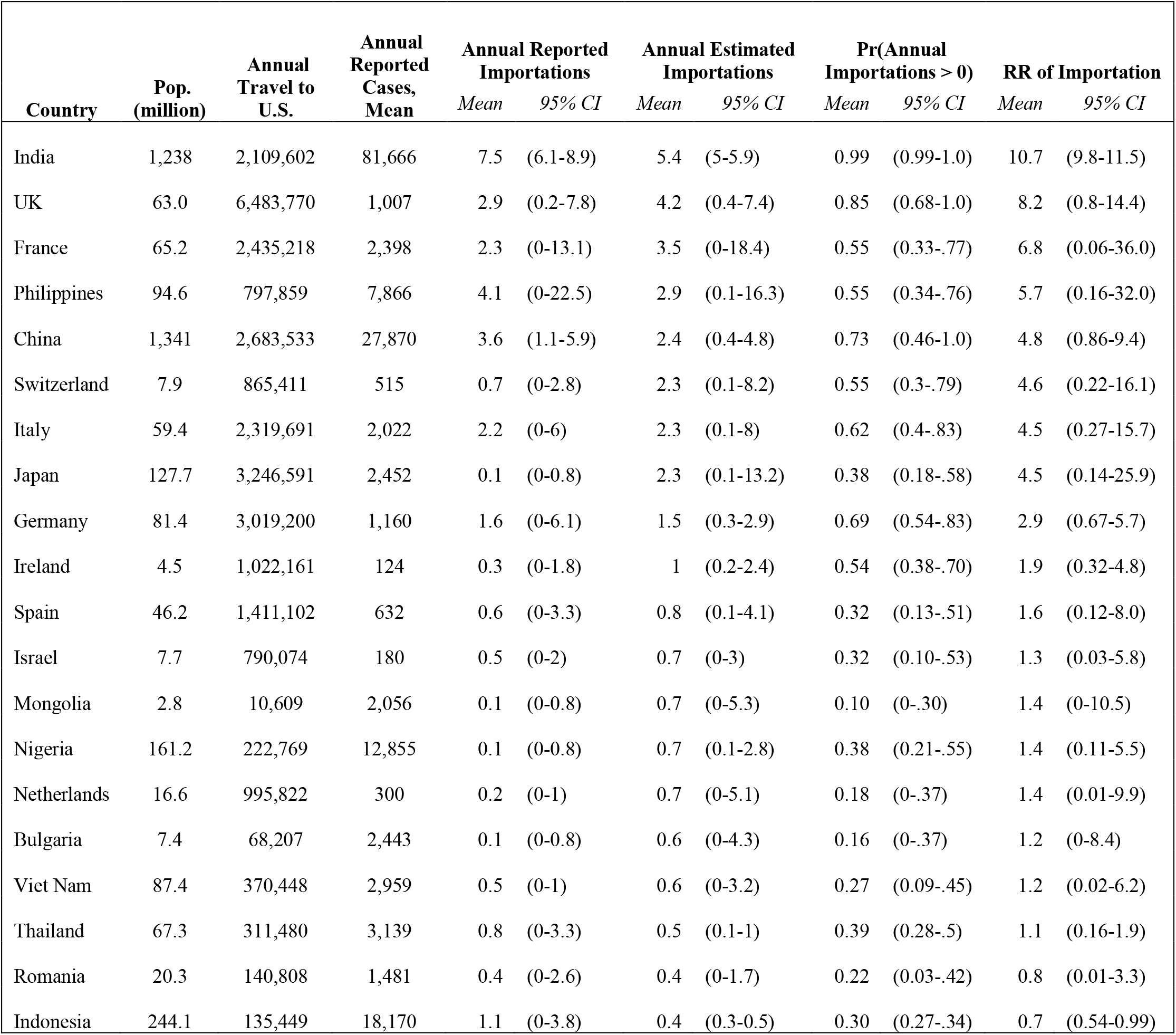
Reported and model estimated mean annual number and risk of measles importations into the United States, from the top 20 source countries, 2006-2015. Relative risk was estimated as the estimated annual importations compared to the median importations across all countries. See *Supplement Table S2* for full table.

**Figure 3.**
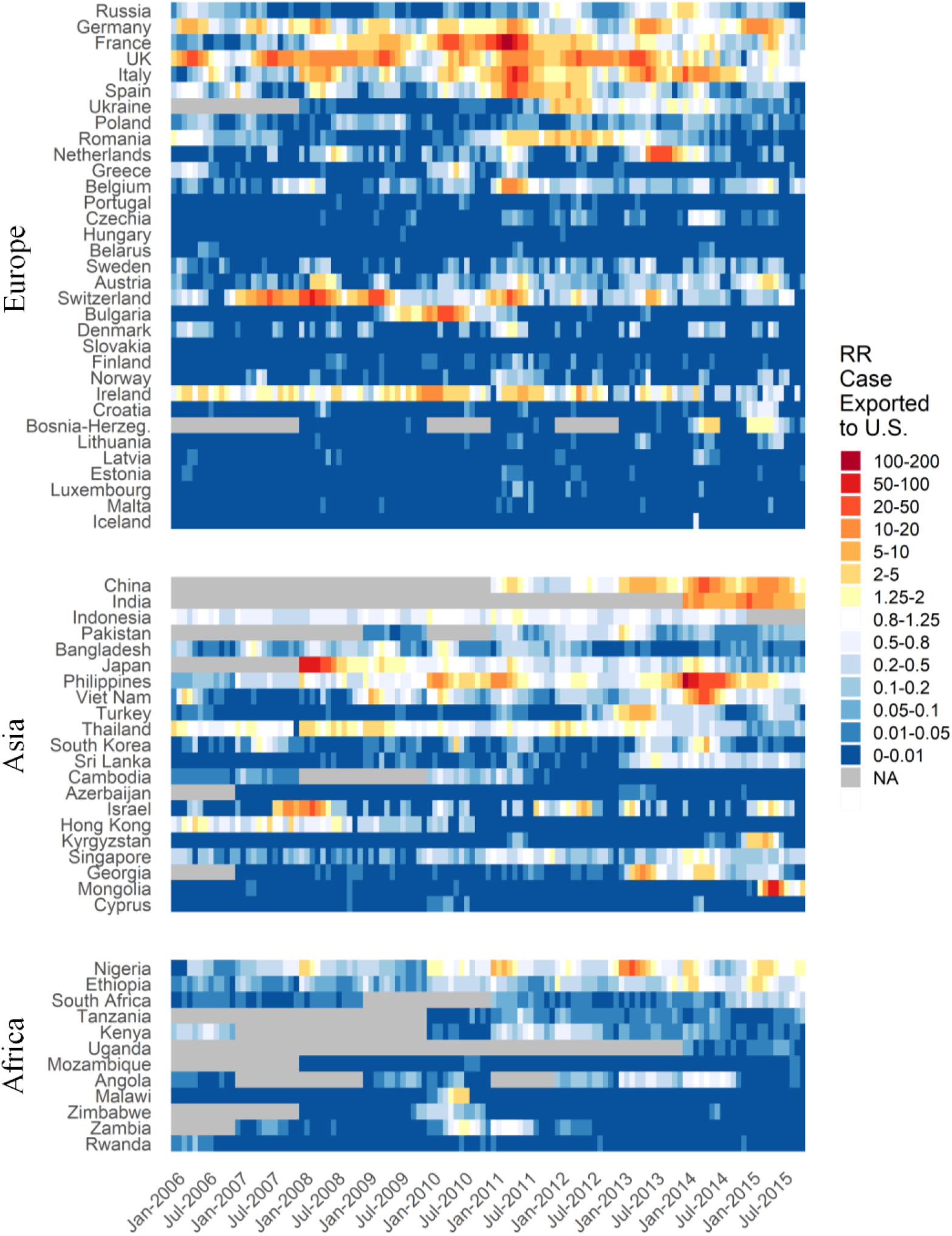
Estimated relative risk of measles importation into the United States by month from 66 countries in Europe, Asia, and Africa January 2006 – December 2015, sorted by country population. Relative risk is calculated against the mean monthly risk of exportation to the United States across all countries and months. Outbreaks in source countries can be clearly identified by the contrasting colors across a country’s risk band (i.e., periods of yellows, oranges, and reds in countries with blues normally). Not all outbreaks denote substantial risk of importation, though contrasts still indicate the outbreak (e.g., Malawi, where a large outbreak occurred in 2010). Other countries can be noted to produce prolonged or continuous importation risk, indicated by long bands of yellow and orange; these are typically countries with endemic measles transmission and high travel volume to the United States (i.e., the UK). Gray bands represent time periods during which importations could not be estimated due to missing measles incidence data from source countries.

### Outbreaks and associated importation risk

The periods of greatest estimated risk during 2006-2015 occurred during the first halves of 2008, 2011, and 2014, corresponding to major outbreaks in Japan and Switzerland (2008), France (2011), and the Philippines (2014) (Figure 3–4). Major outbreaks do not always constitute high risk: while the 2011 outbreak in France (16,350 cases reported) produced 17 reported importations, the larger 2010 outbreak in Bulgaria (23,500 cases reported) produced none (Table S6). While Bulgaria had more cases, annual traveler volume from Bulgaria to the United States was only 3% of that from France. Accounting for these differences in cases, travel, and population, we estimated the importation risk from the French outbreak was 5.2 (95% CI, 1.8-15.2) times as high as from the Bulgarian outbreak, with 23 (95% CI, 20-33) versus 6 (95% CI, 4-11) expected importations into the United States, respectively.

**Figure 4.**
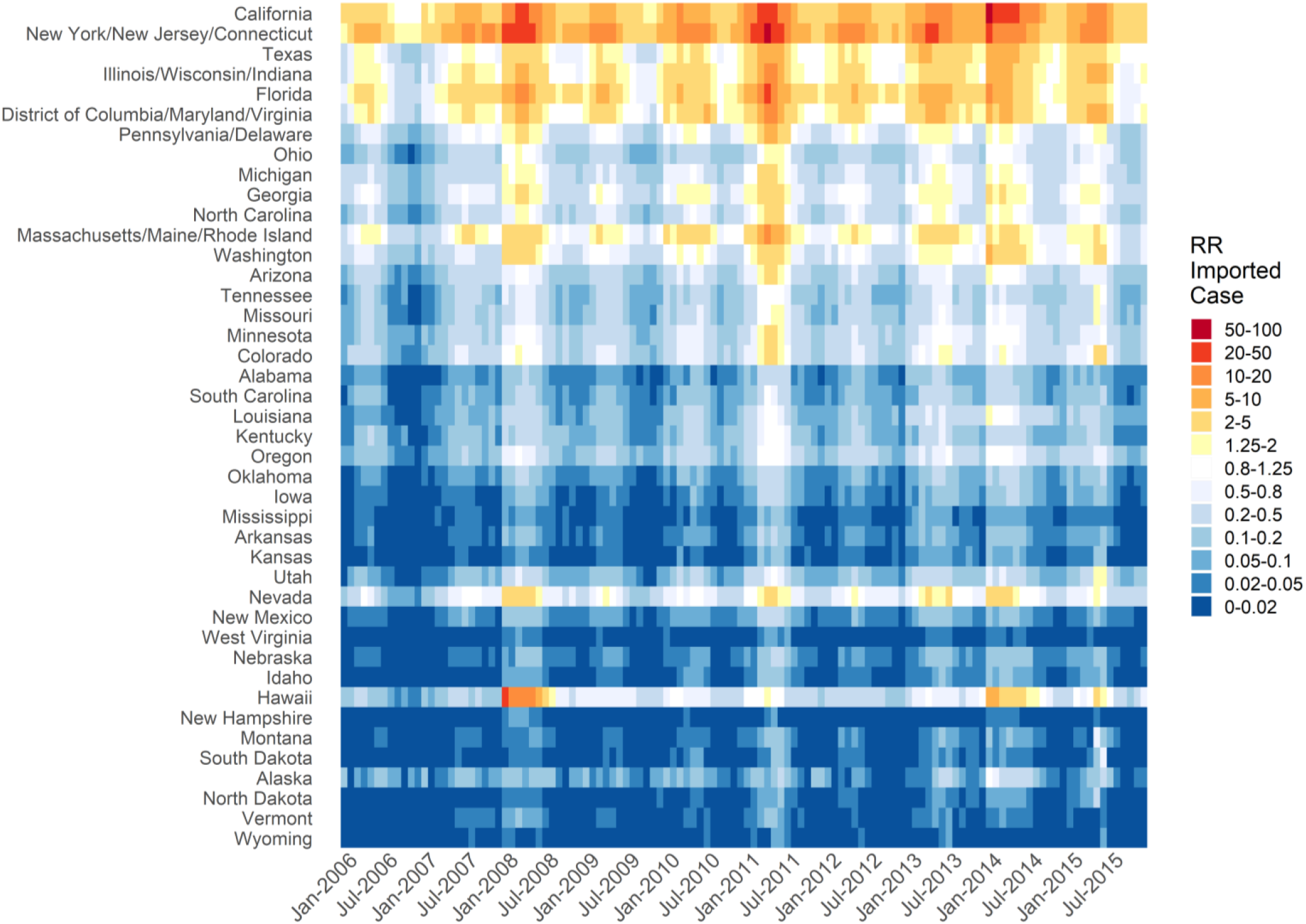
Estimated relative risk of measles importation into the United States by month among U.S. states, January 2006 – December 2015. Relative risk is calculated against the mean monthly risk of importation across all states and all months. States are sorted by population. Periods reflecting outbreaks in countries abroad are reflected by vertical bands of increased relative risk of importation across all states. The periods of highest risk were found during 2008, 2011, and 2014.

### Seasonality and timing of importation risk

Global measles cases (Figure 1) and travel (Figure 2) fluctuate seasonally, driving seasonality in importation risk into the United States, with elevated risk typically occurring in April–July (Figures 3, 5). Measles incidence typically peaks between March and May for Europe and Asia but is more irregular in Africa (Figure S2). In contrast, travel to the United States from all regions peaks in July and August (Figure S3). The temporal alignment of incidence and travel is critical to importation risk. Were measles cases in Europe to peak three months later (in July), when travel between the United States and Europe is at its height, we estimate the number of importations from Europe would increase by approximately 15% (95% CI, 8-32%). For Asia and Africa, this shift in measles timing would increase importations by 4% (95% CI, 3-5%) and 11% (95% CI, 2-12%), respectively.

### Estimated risk of importation, by state

Our model predicted that New York/New Jersey/Connecticut (NY/NJ/CT, combined because of shared airports) and California had the highest risks of importation among U.S. states, experiencing 10.4 (95% CI, 8.5-12.3) and 8.4 (95% CI, 6.7-10.2) times the mean risk across all states during the study period (Figure 5A-B, Table 3). We estimated there were 7.8 (95% CI, 0-40) and 6.2 (95% CI, 0-35) annual importations on average, into these locales, respectively. Risk correlates highly with mean annual state travel volume and population size (Pearson’s *r*=0.88, *r*=0.89). Several states remained at significantly elevated risk after accounting for population size (NY/NJ/CT, California, Hawaii) and travel volume (California, Hawaii; Table 3). Hawaii and West Coast states experienced greater importation risk from Asian countries, while states on the East Coast were more at risk from Europe (Figure 5C). In Florida, 82% of estimated annual importations were associated with travelers originating from European countries, contrasting with only 40% for Washington. For Hawaii, in one year alone (2008), Japan contributed 53% (95% CI, 28-100%) of the risk during the ten-year study period.

**Figure 5.**
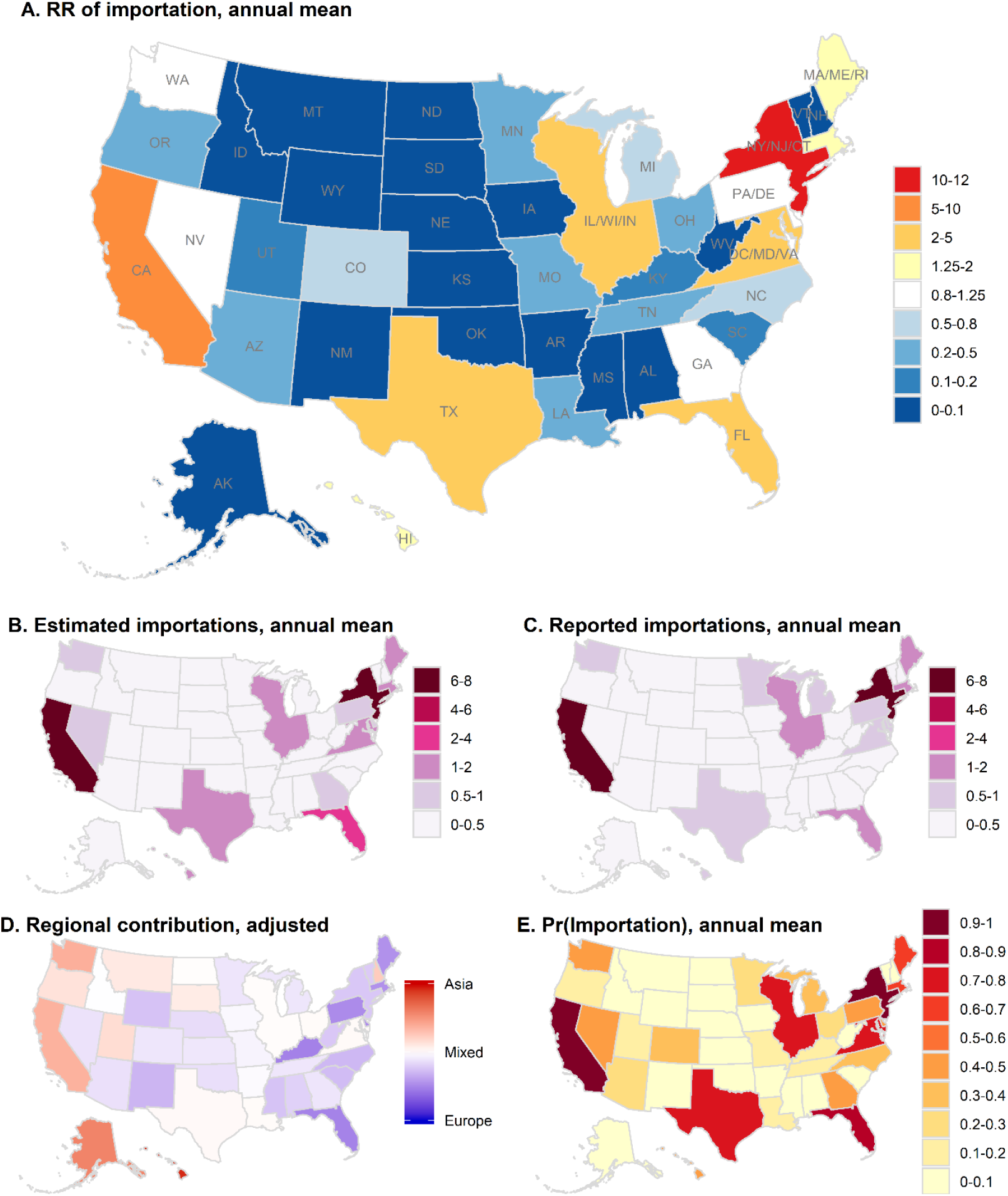
Annual estimated measles importation risk by state or state group within the United States during January 2006 – December 2015. (A) Annual mean importation relative risk by state/state group, calculated as the ratio of the state mean annual estimated number importations, to the mean of state mean annual estimates. (B) Mean annual estimated number of importations by state. (C) Mean annual reported number of importations by state. Only slight differences are evident compared with estimated importations in B. (D) Relative contribution of Asia versus European countries to the risk of importation, adjusted for overall Asia/Europe total ratio of importations (1:1.5 Asia to Europe). Importation risk from Africa was minimal and is not included. (E) Estimated mean annual probability of any importation.

**Table 3.** Reported and model estimated mean annual number and risk of measles importations into the United States, into the top 20 destination states, 2006-2015. Relative risk was estimated as the estimated annual importations compared to the median importations across all states. See *Supplement Table S3* for full table.

| State | Pop.<br>(million) | Annual<br>Travel to<br>U.S. | Annual<br>Reported<br>Importations |  | Annual Estimated<br>Importations |  | Pr(Annual<br>Importations > 0) |  | RR of Importation |  |
| --- | --- | --- | --- | --- | --- | --- | --- | --- | --- | --- |
|  |  |  | Mean | 95% CI | Mean | 95% CI | Mean | 95% CI | Mean | 95% CI |
| New York/<br>New Jersey/Connecticut | 31.8 | 3,375,029 | 6.3 | (3.0-9.0) | 7.8 | (3.3-15.8) | 0.99 | (0.98-1.0) | 10.6 | (4.5-21) |
| California | 37.5 | 6,789,357 | 7.9 | (0.5-20) | 6.2 | (2.2-13.8) | 0.97 | (0.95-1.0) | 8.4 | (3.0-19) |
| Florida | 19.1 | 3,229,180 | 1.4 | (0-3.8) | 2.7 | (1.2-5.3) | 0.88 | (0.82-0.94) | 3.6 | (1.6-7.2) |
| District of Columbia/<br>Maryland/Virginia | 14.5 | 776,860 | 0.6 | (0-1.8) | 2 | (0.7-3.4) | 0.80 | (0.70-0.90) | 2.7 | (1.0-4.6) |
| Illinois/Wisconsin/Indiana | 25.0 | 852,992 | 1.8 | (0-6.8) | 1.8 | (0.7-3.2) | 0.79 | (0.69-0.88) | 2.5 | (0.98-4.4) |
| Texas | 25.4 | 1,716,217 | 1.0 | (0-4.0) | 1.6 | (0.5-2.9) | 0.71 | (0.58-0.84) | 2.1 | (0.68-4.0) |
| Massachusetts/<br>Maine/Rhode Island | 9.0 | 386,829 | 2.0 | (0-8.1) | 1.3 | (0.6-2.6) | 0.67 | (0.58-0.77) | 1.7 | (0.80-3.6) |
| Hawaii | 1.4 | 1,647,029 | 0.7 | (0-4.3) | 1.1 | (0.1-5) | 0.42 | (0.24-0.60) | 1.5 | (0.14-6.8) |
| Washington | 6.8 | 836,760 | 1.0 | (0-3.6) | 0.8 | (0.2-1.5) | 0.49 | (0.37-0.62) | 1.0 | (0.33-2.1) |
| Georgia | 9.7 | 808,274 | 0.2 | (0-1.0) | 0.7 | (0.3-1.2) | 0.48 | (0.39-0.57) | 0.9 | (0.42-1.7) |
| Pennsylvania/Delaware | 13.6 | 173,690 | 0.6 | (0-2.0) | 0.7 | (0.3-1.5) | 0.45 | (0.34-0.55) | 0.9 | (0.37-2.1) |
| Nevada | 2.7 | 740,923 | 0.1 | (0-0.8) | 0.6 | (0.3-1.1) | 0.45 | (0.35-0.55) | 0.9 | (0.34-1.4) |
| Colorado | 5.1 | 461,876 | 0.1 | (0-0.8) | 0.4 | (0.2-0.7) | 0.32 | (0.23-0.40) | 0.5 | (0.22-0.93) |
| Michigan | 9.9 | 590,044 | 0.7 | (0-2.8) | 0.4 | (0.2-0.8) | 0.34 | (0.25-0.43) | 0.6 | (0.26-1.2) |
| North Carolina | 9.6 | 499,399 | 0.2 | (0-1.0) | 0.4 | (0.2-0.9) | 0.33 | (0.24-0.42) | 0.6 | (0.22-1.2) |
| Arizona | 6.4 | 342,301 | 0.4 | (0-1.0) | 0.3 | (0.1-0.6) | 0.25 | (0.18-0.31) | 0.4 | (0.16-0.79) |
| Minnesota | 5.3 | 369,575 | 0.8 | (0-2.0) | 0.3 | (0.1-0.6) | 0.26 | (0.19-0.33) | 0.4 | (0.14-0.79) |
| Ohio | 11.5 | 335,813 | 0.4 | (0-2.6) | 0.3 | (0.1-0.5) | 0.24 | (0.17-0.31) | 0.4 | (0.12-0.66) |
| Louisiana | 4.5 | 152,783 | 0.1 | (0-0.8) | 0.2 | (0-0.4) | 0.14 | (0.08-0.20) | 0.2 | (0.05-0.53) |
| Missouri | 6.0 | 261,379 | 0.5 | (0-2.6) | 0.2 | (0.1-0.4) | 0.20 | (0.14-0.26) | 0.3 | (0.11-0.57) |

### Model Accuracy and Validation

The model’s 95% prediction interval estimates successfully captured the reported number of measles cases imported into the United States for 99% percent of the 7,260 country-months under study (Figure 6). We also assessed its ability to predict the occurrence of at least one importation, which we defined as the probability of at least one imported case being greater than 50%. Sensitivity of this metric was low for both source countries (18%) and U.S. states (18%) and specificity was high (99% for both), indicating that many low-probability importations occur. The positive predictive value was 40% for sources and 49% for states indicating that the 50% probability threshold is a well-calibrated and reliable indicator of risk. Negative predictive value was very high for both (greater than 97%). Model estimates of imported cases correlated highly with numbers of reported imported cases, for both annual and mean country-specific imported cases (*ρ* =0.71 and *ρ* =0.83), while correlating poorly or moderately with travel volume, measles incidence, and population (*ρ*=0.31-0.53, Figure S5-S6). We also compared the full model (with both travel and incidence data) to travel-only or incidence-only models, finding that the full model was significantly more likely to predict the reported importations (likelihood ratio [LR]=44 compared to incidence-only and LR=22 compared to travel-only; Table S5). It should be noted that reported importation cases were not used in any way to fit or calibrate the model, and this performance was obtained completely from reasoning from first principles.

**Figure 6.**
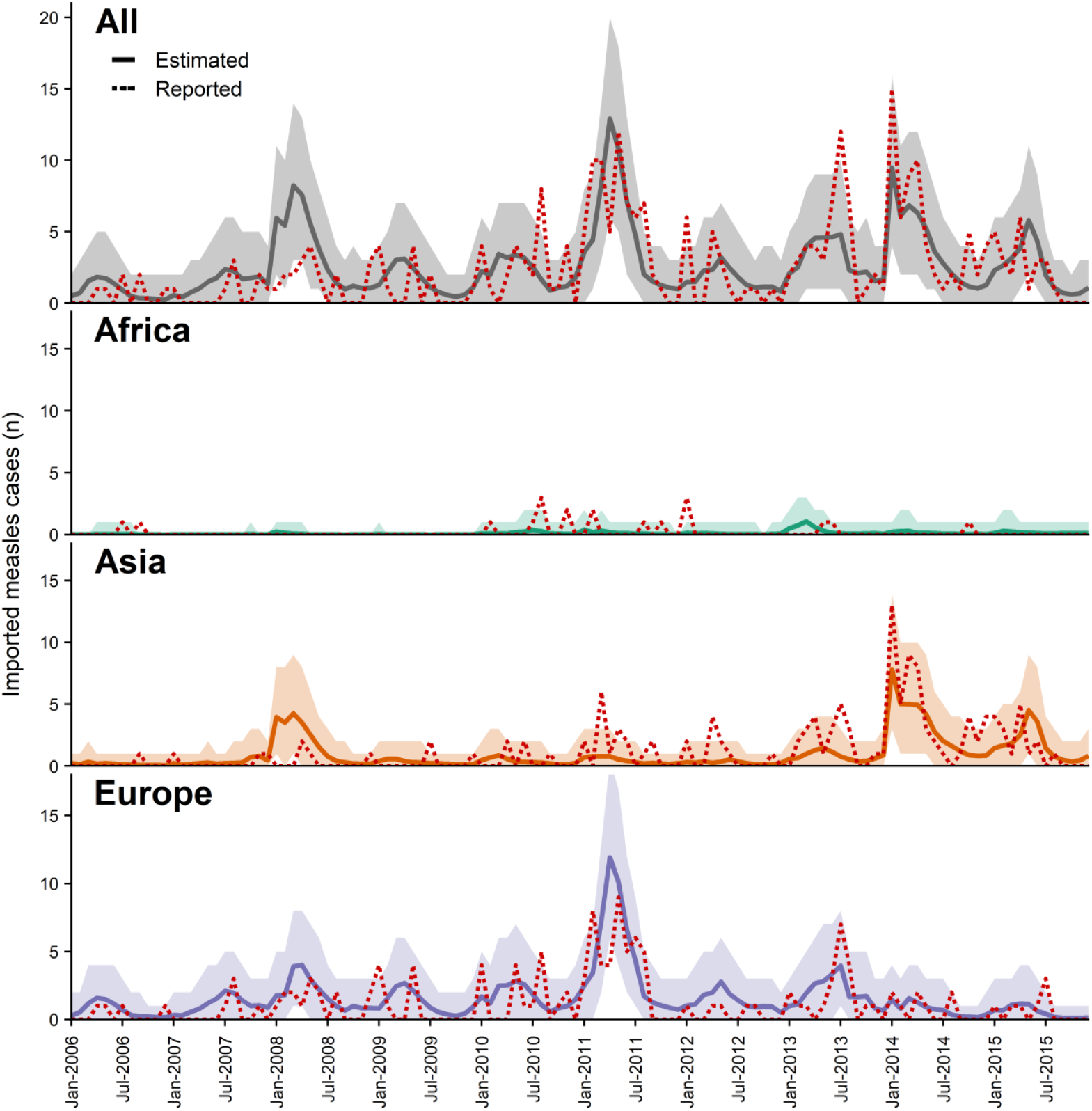
Predicted and reported monthly measles cases imported into the United States, January 2006 – December 2015. Total global and regional monthly measles cases imported into the United States. Solid lines and prediction intervals represent the results from the model using air travel volume and source country measles case counts, and red dotted lines represent the reported measles cases imported into the United States, by month (n=290). We see that overall and regionally, the reported numbers of imported measles cases fall within the aggregated monthly prediction intervals for the majority of months, with 91% of reported cases falling within the 95% prediction interval overall, and 96%, 91%, and 96% for Africa, Asia, and Europe. Accuracy of disaggregated model estimates (per country-month) was significantly higher (99%, 98%, and 99% for Africa, Asia, and Europe, and 99% overall).

### Forecasting measles importations

We conducted a secondary analysis predicting measles importations using forecasted travel data for 2016-2019. Predicted importations were not validated against reported importations, as full details on imported cases were not yet available for this time period. The greatest importation risk occurred in NY/NJ/CT, California, and Florida, associated with travelers originating from both Asia and Europe. The periods of greatest importation risk occurred during March-May 2018, with the Philippines, Italy, France, India, UK, Greece, and Ukraine contributing the most risk, and October 2018-February 2019, from Israel, Ukraine, and Philippines (Figure 7). During this latter period, we estimated that 14.3 (95% CI, 7-22) importations into the United States occurred from Israel, with 5.8 (95% CI, 2-11) of these into NY/NJ/CT. This is a significant divergence from previous risk from Israel, from which there were only 5 reported and 7 (95% CI, 2-12) estimated importations for 2006-2015.

**Figure 7.**
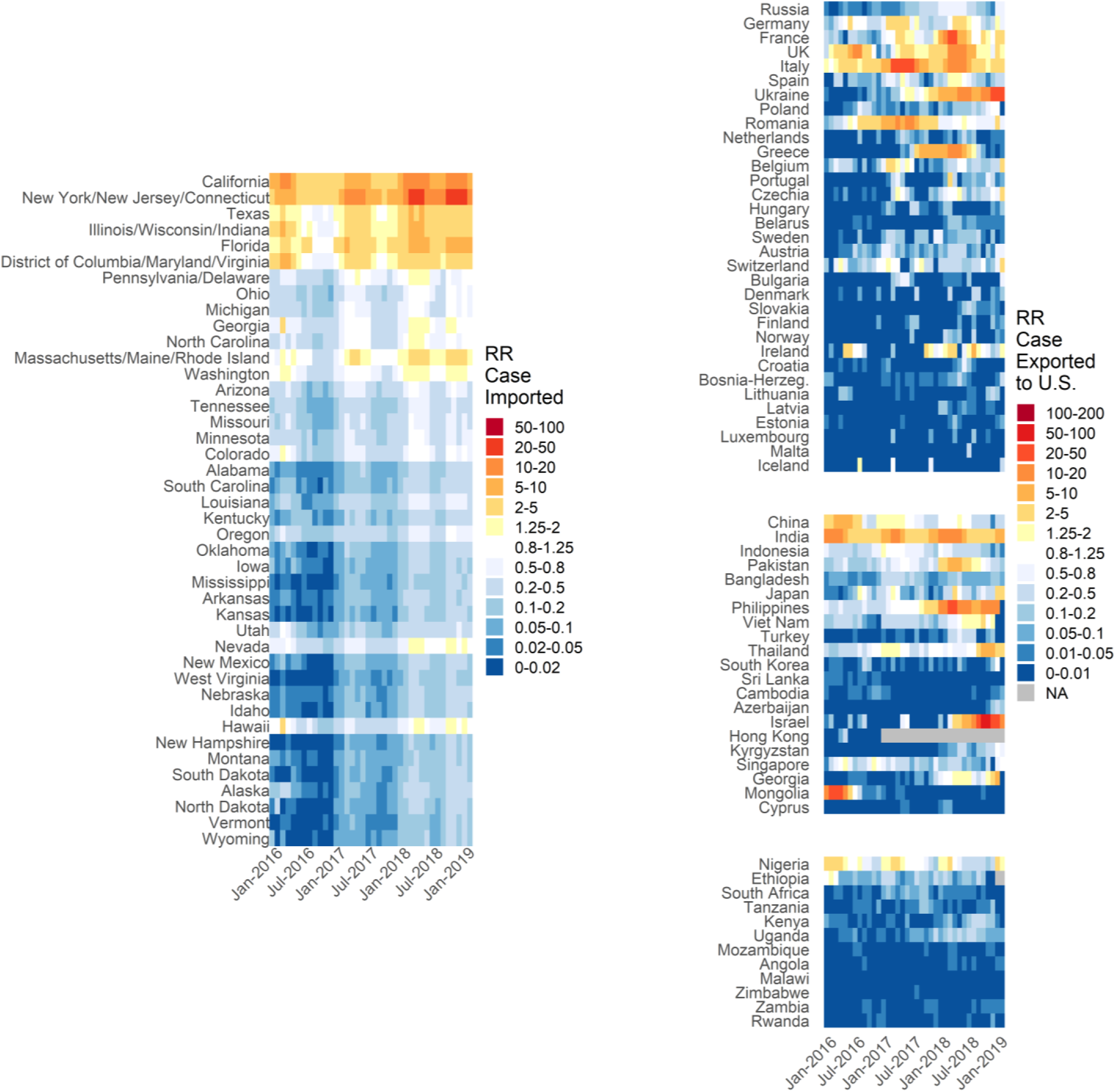
Forecasted estimated relative risk of measles importation by month among U.S. states, January 2016 – February 2019, sorted by state population. Relative risk is calculated against the mean risk of importation across all states and all months. Forecast estimates used reported measles cases and forecasted travel volume. Periods of elevated risk were projected for early and late 2018, associated with outbreaks in the Philippines, across Europe, and in Israel.

## DISCUSSION

Two decades after declaring measles elimination, the United States continues to experience measles outbreaks, reporting a record 1,172 cases during January-August 2019^11,24–26^. This measles incidence is not only a public health challenge for the U.S, but a global problem, with numerous countries at or close to elimination recently experiencing significant outbreaks. Increasing measles incidence and outbreaks can be attributed to insufficient vaccination coverage, at least partly caused by vaccination hesitancy and apathy^27^. With increasing global incidence, the potential for reintroduction of measles virus into countries where the disease had been eliminated, particularly through air travel, poses a significant threat. During 2000-2016, a quarter of measles cases were directly imported into the United States, and the rest resulted from those importations^28^.

As in the United States, recent outbreaks in the Republic of Korea^29^, Israel^30^, Japan^31^, New Zealand^32^, and other countries demonstrate importation of measles virus as a major challenge for control. While herd immunity limits endemic transmission and large-scale outbreaks in the United States, without complete immunity (United States was estimated in 2010 to have 92% overall immunity), potential for transmission from importations remains. This becomes particularly problematic when infected travelers live or interact within clusters of susceptibility, producing even greater potential to spark local transmission^33–35^. To mitigate the risk posed by frequent reintroductions, it becomes critical that we understand when and from where infected travelers will arrive, and where they will go, enabling public health agencies to potentially limit their numbers through public awareness and vaccination and limit transmission through prioritized resources and rapid detection and response.

Previous work has used case and travel data to extrapolate the risk of measles importation. Work by Sarkar et al. recently highlighted the potential to assign risk based on source case and travel data, and how importation can compound outbreak risk produced by non-medical exemptions^36^. Massad et al. estimated the number of imported measles cases into Saudi Arabia during Hajj, emphasizing the risk of transmission and global dissemination following the annual pilgrimage^37^. While these previous studies have demonstrated the value of accounting for both travel and case data, our study is the first to estimate monthly measles importation risk and numbers from international sources into the United States and validate them with detailed data on reported importations collected by the CDC. As we demonstrate, because measles incidence and travel fluctuate over time, correctly accounting for their synchrony through highly time-resolved estimation is critical to appropriately attribute risk. This time-resolution, coupled with measles-specific clinical characteristics (i.e., incubation time, time to characteristic rash), enabled us to accurately project the monthly number of measles importations from specific source countries to specific states in the United States

As expected, we found that importation risk typically concentrates in states/districts with large populations or those with airports that serve as common ports of entry (e.g., New York, California, District of Columbia, and Massachusetts), though some exceptions were found (e.g., Hawaii). We found that two factors (cases of disease in the country of origin and air travel volume) combined to drive importation risk. Periods of elevated importation risk were generally the consequence of large-scale outbreaks (e.g., in France or the Philippines). However, the minimal importation risk produced by large outbreaks in Bulgaria and Malawi demonstrate the necessity of sufficient travel. Overall, both Asia and Europe contributed similar overall importation risk to the United States, with Asia’s greater measles cases countering Europe’s travel volume. Despite massive measles outbreaks and high incidence, Africa contributes only minimal estimated and reported risk, highlighting the critical importance of accounting not only for cases, but also connectivity.

Real-time application of this model demonstrates enormous potential for identifying and responding to emerging risk (Figure 7). In October 2018, eight imported cases from Israel were identified in New York and New Jersey, resulting in outbreaks totaling at least 275 cases largely concentrated among the orthodox Jewish community^38^. Historically, Israel has presented a minimal importation risk to the United States, with only 5 total importations reported from 2006-2015 (we estimated 7). Using projected travel data and real-time surveillance data, our model estimated a dramatic increase in risk, peaking in October 2018, coincident with an outbreak of 4,250 reported in Israel. Of interest, the Israeli outbreak resulted from international travelers importing measles to that country^30^.

The model framework presented here is a parsimonious representation of a much more complex system, and discrepancies between reported and estimated importations partially reflect the model’s simplicity. We do not account for individual-level characteristics, such as vaccination status, or the correlation between travel and infection probabilities—both likely invalid in many populations. Age, for example, is typically associated with infection risk (through susceptibility and contact) and individual likelihood of traveling (e.g. travel purpose, destination)^39,40^. Traveler’s countries of residence, which was not available at the granularity needed and thus not accounted for, may impact the probability of infection: U.S. residents constituted 40% of international travel to the United States, but 62% of imported measles cases (during 2001-2016)^28^. However, the travel data do account for the actual origin country and U.S. destination state/state group, giving confidence to our attribution of risk to locations. Furthermore, while we assume people do not travel once rash has begun, it is possible that some infected individuals do travel with rash or develop rash during travel.

While random individual variability should have minimal impact on accuracy, population-wide characteristics can have a large impact. We saw this during the 2010 Bulgarian outbreak where 90% of cases were among the Roma population and, despite 6 estimated importations, none were reported^22,41^. Similarly, the 2013 Dutch outbreak, which was concentrated (>84%) among spatially-isolated orthodox Protestants, produced only 2 reported importations compared with 7 estimated^42^. Both populations have well-documented histories of insufficient vaccination, are typically socially and spatially isolated, and may be less likely to travel internationally or interact with travelers^41–43^.

This model is also sensitive to data limitations, particularly with reported measles cases (often underestimated and underreported^44–47^) and reported importation cases (*r*equired for validation). For an imported case to be reported, the person must either (1) seek medical care, be diagnosed, then be reported by the provider, or (2) be linked to secondary cases who sought care. Even with highly characteristic measles symptomology and essentially no asymptomatic infections, historically, measles reporting has been incomplete, even in the United States^46–49^. Moreover, foreign visitors might be less likely to seek care, particularly if they depart from the United States early during the infectious period. Importation estimates during a large measles outbreak in Japan during 2007-2008 highlight the challenges of using reported data. While an estimated 4,000-18,000 cases occurred in 2007^50,51^, World Health Organization (WHO) data on reported measles cases include only five cases. This significant underreporting of cases in the country of origin (i.e., Japan) led our model to estimate no importations when ten were reported. In contrast, following national policy changes to require reporting of all measles cases, Japan reported more than 17,000 cases in 2008, from which our model estimated 16 importations. Interestingly, no importations into the United States from Japan were reported during 2008, potentially resulting from changes in vaccination policies, travel advisories, behavior changes, or underreporting of importations^50^. Similarly for Pakistan, reported cases were consistently lower than those found in published sources, contributing to fewer estimated importations than reported^52^.

Despite clear limitations, our simple model does surprisingly well at estimating the number of cases imported into the United States based solely on reasoning from first principles. Similar models simulating yellow fever epidemics^15^, chikungunya virus spread in the Americas^16^, and dengue importation in China^53^ have also demonstrated high accuracy, indicating that this model framework emulates the key processes of disease importation. This accuracy also suggests that some of the more difficult to measure components (e.g., reporting rates, traveler characteristics, population mixing) likely are balanced across most of these countries, most of the time. Nonetheless, a model like this is unlikely to predict rare, but impactful importation events such as that which caused the 2014 outbreak in an Ohio Amish community or the 2017 outbreak in the Minnesota Somali community^34,54^.

For the United States and other governments concerned with importation of diseases like measles, this work highlights two important priorities. First, reducing the risk contributed by resident travelers is essential, accomplished through increased risk awareness^55^ and vaccination coverage^56^ among travelers (particularly high-risk travelers and those from communities with heterogeneous vaccination coverage^57^). Second, this work stresses the tremendous value of information to both domestic biosecurity and global disease control. Our world is highly connected, and infectious diseases are global problems that require global cooperation and efforts, including global disease surveillance systems. The better global systems are at collecting and sharing data, the more timely and accurate those data will be, and the more likely it is that they will be useful in preparedness efforts to prevent outbreaks and protect people. Here we focus on measles importations into the United States, but our methods can readily be applied to other countries and other diseases. Virulent pathogens, like arboviruses, zoonotic coronaviruses, Nipah virus, and highly antibiotic-resistant bacteria, continually emerge around the world and, with the right conditions, can rapidly spread across local and global populations. When this occurs, we need accurate case information and effective tools to guide efforts to prevent and control these disease threats.

## CONCLUSIONS

Until we achieve global eradication, measles virus importations will continue. These importations present a continual risk to spark measles outbreaks in the United States and elsewhere, even after national elimination. With effective tools, like this model, and timely surveillance and travel data, we can identify high-risk destination locations and high-risk source countries in real-time, potentially enabling us to mitigate future biosecurity threats from infectious disease importations through preemptive actions. Increasing and maintaining high routine vaccination coverage, in both the United States and abroad, is the only foolproof means of minimizing the number and the subsequent impact of measles importations.

## METHODS

### Model

Using monthly measles case counts and air travel volume from 66 countries in Europe, Asia, and Africa, we estimated the expected numbers of imported measles cases and the relative risk of measles importation into forty-two U.S. destination states or regions. Our approach was based on previous models of importation for arboviruses.^15,16^ We assumed that importation primarily occurs through air travel, and that it is directly proportional to the number of travelers and the number of cases of infection in the source population. We estimated the monthly number of infected travelers arriving at destination *d* as a binomial process dependent on the number of infected individuals (*I_s,m_*) in source country *s* during month *m*, the duration of infection during which an individual could travel (*D*), and the probability that an individual travels from *s* to *d* and is detected in *d* during month *m* (*p_s_,_d_,_m_*). This is estimated as follows:

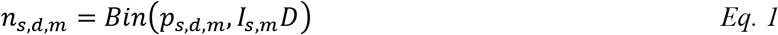

Using the monthly number of travelers from *s* to *d* (*T_s,d,_*_m_), the total population size of *s* (*N_s,m_*), and the number of days per month (*δ_m_*), we approximated *p_s,d,m_*:

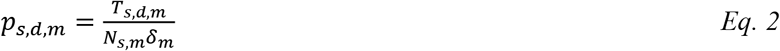

We performed 10,000 stochastic simulations, sampling *D* from a distribution for each simulation, and calculating *n_s,d,m_* for each *s, d*, and *m* combination.

### Data

#### Source country measles case and population data

Monthly reported numbers of measles cases are publicly available from the World Health Organization and the European Centers for Disease Control. These data were checked against country and regional reports where possible to verify accuracy. Annual case counts from monthly data were checked against reported annual measles cases available from the WHO^21^. Where annual totals differed by more than 25%, monthly data were considered invalid and excluded. Monthly case data were missing for multiple years for several countries. Where data were deemed invalid or missing, these country/year combinations were excluded from the analysis (e.g., India 2006-2013). Population data are publicly available from the United Nations World Population Prospect data^58^.

#### Travel data

Summary travel data are proprietary data from Diio and OAG in two batches^59, 60^. First, data were collected for 28 European countries for 2006-2015 from Diio in October 2016. Data for the additional 38 countries – 21 Asian, 12 African, and 5 European countries (Belarus, Bosnia-Herzegovina, Croatia, Switzerland, Ukraine) for 2010-2015 were collected from OAG in July 2017. These data are modeled estimates of trip numbers based on ticket sales and reporting from carriers and include locations of initial departure and final destination. As international airports like O’Hare International Airport in Chicago or John F. Kennedy International Airport in New York also serve nearby states, we grouped several of these states together to limit bias, though this likely does not fully resolve this bias. To provide model results for all 66 countries for a ten-year span from January 2006 – December 2015, we imputed missing travel data from OAG using the available data and regionally-aggregated monthly travel data from the U.S. National Travel and Tourism Office^61^ (Table S8). The imputation model was validated using the 28-country European travel data set, using leave-one-out methods (see *Supplement for more details*).

#### Measles importations to the United States

To validate the model, we use data on the reported number of cases of measles virus infection imported into the United States, which were provided by the U.S. Centers for Disease Control and Prevention. These data include the month and state of detection, and country of origin or travel. Reported importations that originated from countries other than the 66 included or during time periods where travel or measles case data were not available were excluded to ensure comparability with model results (n=112). Among the excluded reported importations, 50 originated in India, occurring during 2006-2013, during which monthly measles case data are not available for India. Because this is such a large portion of the reported importations (12%), we opted to examine the annual means of importation and risk, rather than the ten-year totals for the full study period, whereby annual means were estimated across years for which data were complete. Because India reported relatively consistent annual measles case counts (median=32,546; IQR=27,440-43,480), the estimated mean annual cases and importations for India should not change substantially were data for 2006-2013 included. We also should note that the numbers of importations that we report do not fully concur with those previously published^28^, as individuals reporting multiple potential countries of infection (n=22) were counted as an importation from each reported country.

#### Disease Parameters

The duration during which an infected individual could travel post-infection (*D*). For measles, we assumed individuals do not travel once the rash develops. Based on previous estimates, we quantify *D* as the sum of a normally-distributed incubation period (mean=12.5 days, 95% CI=11.8- 13.3) and a uniformly-distributed time to rash (2-3 days)^62^.

Primary analysis data were restricted to January 2006 through December 2015, the period for which data were complete for all sources.

#### Forecasting Analysis

To demonstrate our ability to forecast the risk of measles importation, especially in real-time, we repeated the primary analysis for January 2016 through February 2019, using a combination of reported measles case data and forecasted travel data for all 66 countries. These forecasted travel data were generated using the same model constructed to impute missing travel volume data.

## Model validation

Model performance was evaluated using Pearson’s correlation between the estimated importations and risk, and reported importation cases, by year and by month. We further validated the model through comparison of fits to reported importation data with those of probability models that excluded data (i.e., excluded travel data, case data, and both; s*ee Supplement*).

## Relative risk of importation

Using the simulation results for the absolute number of importations, we also estimated the relative risks of importation. There are a multitude of factors that can directly influence the absolute magnitude of importations, including individual-level characteristics of travelers and the accuracy of reported measles cases. While we cannot directly account for all these factors, assuming they are relatively consistent across destinations and sources, we can use the relative risk (*R*R) of importation as a metric to compare risk between destinations and sources. We estimate the RR of importation as the ratio of the estimated number of imported cases to the mean number of importations for that simulation across all time periods and sources or destinations as the reference. Thus, for each simulation, a single reference value is established across all countries and time periods. The mean RR and confidence intervals across the 10,000 simulations are estimated for each destination/time or source/time combination.

All analyses were performed using R 3.5.2 and RStudio 1.1.463.

## Data Availability

These analyses require three primary sources of data: reported measles cases from source locations, travel volume data, and population data. Reported monthly measles cases are available publicly from the World Health Organization (https://www.who.int/immunization/monitoring_surveillance/burden/vpd/surveillance_type/active/measles_monthlydata/en/). Population data are publicly available from the United Nations World Population Prospects data (https://population.un.org/wpp/). Monthly air travel are primarily from OAG (OAG Aviation Worldwide, https://www.oag.com/). These data were provided under contract and cannot be provided by the authors. Additionally, data on reported measles importation were used for validation. These data were provided by the U.S. Centers for Disease Control and Prevention and are not currently available to the public.

## Acknowledgments

This work was supported by the Bill & Melinda Gated Foundation [S.A.T. and J.L., grant OPP1094793]. The air passenger data used in this study were purchased from Diio, LLC and OAG Aviation Worldwide Ltd. These data were used under license for the current study and so are not publicly available. The authors are available to share the air passenger data upon reasonable request from OAG and with the permission of OAG Aviation Worldwide Ltd. The authors no longer have access to Diio, LLC data. For researchers, the opportunity to access the data would be through the procurement of Diio Mi FMg. Please contact Diio’s Senior Vice President, Jordan Kayloe, at.

The opinions expressed by authors contributing to this journal do not necessarily reflect the opinions of the Centers for Disease Control and Prevention or the institutions with which the authors are affiliated.Author information

Affiliations:

*Johns Hopkins Bloomberg School of Public Health, Baltimore, MD, 21205, USA*

Shaun A. Truelove, Justin Lessler

*Biomedical Advanced Research and Development Authority, U.S. Department of Health & Human Services, Washington, DC, 20201, USA*

Luis Mier-y-Teran-Romero

*Division of Vector-Borne Diseases, Centers for Disease Control and Prevention, Atlanta, GA, 30333, USA*

Allison Taylor Walker, Andre Berro

*Division of Global Migration and Quarantine, Centers for Disease Control and Prevention, Atlanta, GA, 30333, USA*

Paul Gastanaduy

*Division of Viral Diseases, Centers for Disease Control and Prevention, San Juan, Puerto Rico, 00920, USA*

Michael A. Johansson

*Center for Communicable Disease Dynamics, Harvard TH Chan School of Public Health, Boston, MA, 02115, USA*

Michael A. Johansson

Contributions:

S.A.T., L.M.T.R, J.L., and M.A.J. conceived and designed the project. S.A.T. drafted the manuscript.

S.A.T. and L.M.T.R conducted the analyses. S.A.T., L.M.T.R, A.T.W., P.G., A.B., and M.A.J. contributed to data acquisition. All authors discussed the results, contributed critical revisions, and approved the final manuscript.

## References

1. International tourism, number of arrivals | Data. http://data.worldbank.org/indicator/ST.INT.ARVL?end=2015&start=1995&view=map.

2. Monthly Departures to International Destinations. http://travel.trade.gov/view/m-2016-O-001/index.html.

3. Summary of probable SARS cases with onset of illness from 1 November 2002 to 31 July 2003. World Health Organization http://www.who.int/csr/sars/country/table2004_04_21/en/ (2003).

4. Dawood, F. S. et al. Estimated global mortality associated with the first 12 months of 2009 pandemic influenza A H1N1 virus circulation: a modelling study. Lancet Infect. Dis. 12, 687–695 (2012).

5. Forgione, M. MERS health warning signs go up at LAX and other U.S. airports. Los Angeles Times (2014).

6. Thomas, H. L. et al. Enhanced MERS Coronavirus Surveillance of Travelers from the Middle East to England - Volume 20, Number 9—September 2014 - Emerging Infectious Diseases journal - CDC. doi:10.3201/eid2009.140817.

7. Ebola screening begins at Heathrow. BBC News (2014).

8. Cohen, N. J. Travel and Border Health Measures to Prevent the International Spread of Ebola. MMWR Suppl. 65, (2016).

9. Poletto, C., Boelle, P.-Y. & Colizza, V. Risk of MERS importation and onward transmission: a systematic review and analysis of cases reported to WHO. Bmc Infect. Dis. 16, 448 (2016).

10. Vonnahme, L. A., Jungerman, M. R., Gulati, R. K., Illig, P. & Alvarado-Ramy, F. US Federal Travel Restrictions for Persons with Higher-Risk Exposures to Communicable Diseases of Public Health Concern. Emerg. Infect. Dis. 23, (2017).

11. Orenstein, W. A., Samuel, K. L. & Hinman, A. R. Summary and Conclusions: Measles Elimination Meeting, 16–17 March 2000. J. Infect. Dis. 189, S43-S47 (2004).

12. Parker Fiebelkorn, A. et al. Measles in the United States during the Postelimination Era. J. Infect. Dis. 202, 1520-1528 (2010).

13. Fiebelkorn, A. P. et al. A Comparison of Postelimination Measles Epidemiology in the United States, 2009-2014 Versus 2001–2008. J. Pediatr. Infect. Dis. Soc. 6, 40-48 (2017).

14. Data Access - Vital Statistics Online. https://www.cdc.gov/nchs/data_access/Vitalstatsonline.htm.

15. Johansson, M. A. et al. Assessing the Risk of International Spread of Yellow Fever Virus: A Mathematical Analysis of an Urban Outbreak in Asunción, 2008. Am. J. Trop. Med. Hyg. 86, 349–358 (2012).

16. Johansson, M. A., Powers, A. M., Pesik, N., Cohen, N. J. & Staples, J. E. Nowcasting the Spread of Chikungunya Virus in the Americas. PLoS ONE 9, (2014).

17. Minetti, A. et al. Lessons and Challenges for Measles Control from Unexpected Large Outbreak, Malawi. Emerg. Infect. Dis. 19, 202–209 (2013).

18. Ibrahim, B. S., Mohammed, Y., Usman, R., Abubakar, A. & Nguku, P. Burden and Trend of Measles in Nigeria: Five-year Review Case-base Surveillance Data. Online J. Public Health Inform. 10, (2018).

19. Kidd, S. et al. Measles outbreak in Burkina Faso, 2009: A case-control study to determine risk factors and estimate vaccine effectiveness. Vaccine 30, 5000–5008 (2012).

20. Mpabalwani, M. E. et al. The 2010 - 2011 measles outbreak in Zambia: Challenges and lessons learnt for future action. East Afr. J. Public Health 10, 265–273-273 (2013).

21. World Health Organization. Measles and Rubella Surveillance Data. WHO http://www.who.int/immunization/monitoring_surveillance/burden/vpd/surveillance_type/active/measles_monthlydata/en/.

22. Muscat, M. et al. The measles outbreak in Bulgaria, 2009-2011: An epidemiological assessment and lessons learnt. Eurosurveillance 21, 30152 (2016).

23. Al, D. A. et. Measles Elimination Efforts and 2008-2011 Outbreak, France - Volume 19, Number 3—March 2013 - Emerging Infectious Disease journal - CDC. doi:10.3201/eid1903.121360.

24. Region of the Americas is declared free of measles. Pan American Health Organization / World Health Organization http://www.paho.org/hq/index.php?option=com_content&view=article&id=12528%3Aregion-americas-declared-free-measles&catid=740%3Apress-releases&Itemid=1926&lang=en (2016).

25. Patel, M. Increase in Measles Cases — United States, January 1-April 26, 2019. MMWR Morb. Mortal. Wkly. Rep. 68, (2019).

26. CDC. Measles Cases and Outbreaks. Centers for Disease Control and Prevention https://www.cdc.gov/measles/cases-outbreaks.html (2019).

27. Health, T. L. C., & A. Vaccine hesitancy: a generation at risk. Lancet Child Adolesc. Health 3, 281 (2019).

28. Lee, A. D., Clemmons, N. S., Patel, M. & Gastañaduy, P. A. International Importations of Measles Virus into the United States During the Postelimination Era, 2001–2016. J. Infect. Dis. 219, 1616-1623 (2019).

29. Yang, T. U. et al. Resurgence of measles in a country of elimination: interim assessment and current control measures in the Republic of Korea in early 2014. Int. J. Infect. Dis. 33, 12–14 (2015).

30. Measles. Vaccines for Babies and Children https://www.health.gov.il/English/Topics/Pregnancy/Vaccination_of_infants/Pages/measles.aspx.

31. Takahashi, T. et al. Ongoing increase in measles cases following importations, Japan, March 2014: times of challenge and opportunity. West. Pac. Surveill. Response J. WPSAR 5, 31–33 (2014).

32. Hayman, D. T. S. et al. Global importation and population risk factors for measles in New Zealand: a case study for highly immunized populations. Epidemiol. Infect. 145, 1875-1885 (2017).

33. Truelove, S. A. et al. Characterizing the impact of spatial clustering of susceptibility for measles elimination. Vaccine 37, 732–741 (2019).

34. Gastañaduy, P. A. et al. A Measles Outbreak in an Underimmunized Amish Community in Ohio. N. Engl. J. Med. 375, 1343-1354 (2016).

35. Centers for Disease Control and Prevention. Import-Associated Measles Outbreak — Indiana, May--June 2005. Morb. Mortal. Wkly. Rep. 54, 1073–1075 (2005).

36. Sarkar, S., Zlojutro, A., Khan, K. & Gardner, L. Measles resurgence in the USA: how international travel compounds vaccine resistance. Lancet Infect. Dis. 19, 684–686 (2019).

37. Massad, E., Wilder-Smith, A. B., Wilder-Smith, A. & Memish, Z. A. Modelling the importation risk of measles during the Hajj. Lancet Infect. Dis. 19, 806 (2019).

38. McDonald, R. Notes from the Field: Measles Outbreaks from Imported Cases in Orthodox Jewish Communities — New York and New Jersey, 2018-2019. MMWR Morb. Mortal. Wkly. Rep. 68, (2019).

39. Mossong, J. et al. Social Contacts and Mixing Patterns Relevant to the Spread of Infectious Diseases. PLoS Med 5, e74 (2008).

40. Read, J. M. et al. Social mixing patterns in rural and urban areas of southern China. Proc. R. Soc. B Biol. Sci. 281, (2014).

41. Vassilev, R. The Roma of Bulgaria: A pariah minority. Glob. Rev. Ethnopolitics 3, 40–51 (2004).

42. Woudenberg, T. et al. Large measles epidemic in the Netherlands, May 2013 to March 2014: changing epidemiology. Eurosurveillance 22, (2017).

43. Rechel, B., Blackburn, C. M., Spencer, N. J. & Rechel, B. Access to health care for Roma children in Central and Eastern Europe: findings from a qualitative study in Bulgaria. Int. J. Equity Health 8, 24 (2009).

44. Takahashi, H. & Saito, H. Measles Exportation From Japan to the United States, 1994 to 2006. J. Travel Med. 15, 82–86 (2008).

45. van Isterdael, C. E. D. et al. Measles incidence estimations based on the notification by general practitioners were suboptimal. J. Clin. Epidemiol. 57, 633–637 (2004).

46. Woudenberg, T. et al. The tip of the iceberg: incompleteness of measles reporting during a large outbreak in The Netherlands in 2013-2014. Epidemiol. Infect. 1–7 (undefined/ed) doi:10.1017/S0950268818002698.

47. Odega, C. C., Fatiregun, A. A. & Osagbemi, G. K. Completeness of suspected measles reporting in a southern district of Nigeria. Public Health 124, 24–27 (2010).

48. Harpaz, R. Completeness of measles case reporting: review of estimates for the United States. J. Infect. Dis. 189 Suppl 1, S185–190 (2004).

49. Davis, S. F. et al. Reporting efficiency during a measles outbreak in New York City, 1991. Am. J. Public Health 83, 1011–1015 (1993).

50. Nakayama, T. Vaccine chronicle in Japan. J. Infect. Chemother. 19, 787–798 (2013).

51. Progress Toward Measles Elimination — Japan, 1999--2008. https://www.cdc.gov/mmwr/preview/mmwrhtml/mm5738a5.htm.

52. Kazi, A. N. Measles epidemic exposes inadequate vaccination coverage in Pakistan. BMJ 346, f245 (2013).

53. Lai, S. et al. Seasonal and interannual risks of dengue introduction from South-East Asia into China, 2005-2015. PLoS Negl. Trop. Dis. 12, e0006743 (2018).

54. Hall, V. et al. Measles Outbreak — Minnesota April-May 2017. MMWR Morb. Mortal. Wkly. Rep. 66, 713–717 (2017).

55. CDC. Don’t Bring Measles Home. Centers for Disease Control and Prevention http://www.cdc.gov/features/measlesinternationaltravel/ (2019).

56. Hyle, E. P. et al. Missed Opportunities for Measles, Mumps, Rubella Vaccination Among Departing U.S. Adult Travelers Receiving Pretravel Health Consultations. Ann. Intern. Med. (2017) doi:10.7326/M16-2249.

57. Hyle, E. P. et al. The Clinical Impact and Cost-effectiveness of Measles-Mumps-Rubella Vaccination to Prevent Measles Importations Among International Travelers From the United States. Clin. Infect. Dis. 69, 306–315 (2019).

58. World Population Prospects - Population Division - United Nations. https://population.un.org/wpp/.

59. Diio. Data In, Intelligence Out, Market Intelligence, Fares and Market Sizes, Global LLC. https://www.diio.net/.

60. OAG Aviation Worldwide Ltd. OAG Traffic Analyser, Version 2.0. https://analytics.oag.com/analyser-client/home (2018).

61. Monthly Tourism Statistics - Overseas Arrivals to the U.S. https://travel.trade.gov/research/monthly/arrivals/index.asp.

62. Lessler, J. et al. Incubation periods of acute respiratory viral infections: a systematic review. Lancet Infect. Dis. 9, 291–300 (2009).

